# Comparing the performance of risk stratification scores in Brugada syndrome: a multi-centre study

**DOI:** 10.1101/2021.11.09.21266130

**Authors:** Sharen Lee, Jiandong Zhou, George Bazoukis, Konstantinos P Letsas, Tong Liu, Wing Tak Wong, Ian Chi Kei Wong, Ngai Shing Mok, Chloe Mak, Qingpeng Zhang, Gary Tse

## Abstract

**Introduction:** The management of Brugada Syndrome (BrS) patients at intermediate risk of arrhythmic events remains controversial. The present study evaluated the predictive performance of different risk scores in an Asian BrS population and its intermediate risk subgroup.

**Methods:** This is a retrospective territory-wide cohort study of consecutive patients diagnosed with BrS from January 1^st^, 1997 to June 20^th^, 2020 in Hong Kong. The primary outcome is sustained ventricular tachyarrhythmias. A novel predictive score was developed. Machine learning-based nearest neighbor and Gaussian Naïve Bayes models were also developed. The area under the receiver operator characteristic (ROC) curve (AUC) was compared between the different scores.

**Results:** The cohort consists of 548 consecutive BrS patients (7% female, age at diagnosis: 50±16 years old, follow-up duration: 84±55 months). For risk stratification in the whole BrS cohort, the score developed by Sieira et al. showed the best performance with an AUC of 0.805, followed by the Shanghai score (0.698), and the scores by Okamura *et al*. (0.667), Delise *et al*. (0.661), Letsas *et al*. (0.656) and Honarbakhsh *et al*. (0.592). A novel risk score was developed based on variables and weighting from the best performing score (the Sieira score), with the inclusion of additional variables significant on univariable Cox regression (arrhythmias other than ventricular tachyarrhythmias, early repolarization pattern in the peripheral leads, aVR sign, S-wave in lead I and QTc ≥436 ms). This score has the highest AUC of 0.855 (95% CI: 0.808-0.901). The Gaussian Naïve Bayes model demonstrated the best performance (AUC: 0.97) compared to logistic regression and nearest neighbor models.

**Conclusion:** The inclusion of investigation results and more complex models are needed to improve the predictive performance of risk scores in the intermediate risk BrS population.

## Introduction

Brugada Syndrome (BrS) is an ion channelopathy with a characteristic electrocardiographic (ECG) pattern (BrP) of ST-elevation followed by either a coved-shaped (type 1) or saddle-shaped (type 2) slope. This disease predisposes affected patients to an increased risk of sudden cardiac death (SCD) due to sustained ventricular tachycardia/fibrillation (VT/VF) in the absence of overt structural abnormalities. Therefore, the stratification of VT/VF/SCD risk in BrS patients is critical to the management of BrS. Although BrS has a higher prevalence in Asia, a large proportion of existing research was based on registries that include mostly Caucasian subjects. (1-4) As a result, the VT/VF/SCD risk stratification tools derived were also largely based on the Western population. (5,6)

Intermediate risk refers to the presence of risk factors suggestive of high and low risks, such as an asymptomatic patient presenting with spontaneous type 1 BrP. (7) Whilst it is clear that high risk patients should be referred for implantable cardioverter-defibrillator implantation, and low risk patients should be monitored regularly, it is the management of these intermediate risk patients that remains controversial (8). Recently, Probst *et al*. evaluated the predictive value of the Shanghai and Sieira score against intermediate risk BrS patients in the largest cohort of BrS patients to date and concluded that risk scores could not stratify the arrhythmic risk in this subpopulation. (7) However, other existing risk scores were not evaluated, with the Shanghai score not designed to be a prognostic tool. In addition, the Asian population was not assessed despite the greater prevalence of BrS in Asia. Therefore, the present study aims to evaluate the predictive performance of different risk scores in the overall Asian BrS population and its intermediate risk subpopulation, thus examine the applicability of simple risk scores in a clinical setting.

## Method

### Patient Cohort and Data Collection

Ethics approval for the present study was obtained from The Joint Chinese University of Hong Kong – New Territories East Cluster Clinical Research Ethics Committee. The present cohort consists of consecutive patients diagnosed with BrS from January 1^st^, 1997 to June 20^th^, 2020 at centers managed in the Hong Kong public sector. The BrS diagnosis was confirmed by G.T. and N.S.M. after reviewing the relevant case notes and ECGs of the patients based on the Expert Consensus Statement proposed in 2017. (9)

The following clinical data were extracted: 1) sex; 2) presentation at diagnosis: age, type of BrP, symptom (asymptomatic, syncope, or VT/VF), fever-induced BrP; 3) performance and results of drug challenge test, electrophysiological study (EPS) and genetic test; 4) family history of SCD and BrS; 5) follow-up: duration, the occurrence of VT/VF and time from diagnosis if present, mortality and cause if relevant, presence of type 1 BrP; 6) baseline concomitant presence of other arrhythmias (sick sinus syndrome, bradycardia, atrioventricular block, supraventricular tachycardia (SVT), supraventricular ectopic beats (SVE), atrial fibrillation (AF), atrial tachycardia (AT), or atrial flutter). An asymptomatic presentation is defined as the absence of syncope and VT/VF. The evolution of BrP refers to the presentation of other types of BrP other than the type presented at diagnosis.

Automatically measured baseline ECG indices were extracted: 1) heart rate; 2) PR interval; 3) QRS interval; 4) QT and QTc interval; 5) P, QRS and T wave axis. These ECG parameters were averaged across the 12 leads. In addition, G.T. and G.B. identified the following ECG features: 1) early repolarization in peripheral leads; 2) aVR sign (ST elevation in lead aVR); 3) significant S wave in lead 1 (S wave > 0.1mV or >40ms); 4) fragmented QRS. The aforementioned ECG indices were extracted given their potential in reflecting BrS-related ECG changes, thus have potential risk stratification value. (10,11)

### Outcomes, Statistical Analysis, and Machine Learning Models

The primary outcome is sustained VT/VF occurring during follow-up. This was obtained from case notes by the physicians during inpatient or outpatient encounteres, and/or implantable cardioverter-defibrillator documentation where available. Continuous variables were reported as mean (standard deviation), whilst discrete variables were reported as total count (percentage). To identify predictors of the primary outcome, univariable Cox regression was performed. Findings from the drug challenge test and EPS were not included in the model since they were not universally performed. Significant univariable predictors were used as inputs for multivariable Cox regression. Significant predictors from both the univariable and multivariable models were selected to develop predictive scores separately. For continuous variables, cut-off values were identified using the Liu method. The HR and 95% confidence interval (CI) were reported. The weighting of each parameter was adopted from the hazard ratio (HR) calculated from the results of Cox regression.

To evaluate the predictive power of the devised scores against existing scores, the area under the receiver-operator-characteristic curve (AUC) and its 95% CI were generated. The existing scores evaluated are summarized in **Supplementary Table 1**. (6,10-14) In order to evaluate the predictive value of the scores against patients of intermediate risk, the calculation of AUC is repeated after the removal of patients scoring the first and fourth quartile for each score. Random survival forest (RSF) was applied to identify the importance ranking amongst the significant univariable predictors. The importance ranking is measured by the minimal depth and variable importance. A smaller minimal depth means the variable is more important since the variable splits the data further away from the terminal node, whilst higher variable importance refers to a greater change in prediction error when the variable of interest is absent. Statistical significance was defined as P-value <0.05.

In addition, machine learning models were developed, including nearest neighbor model and Gaussian Naïve Bayes model, to predict sustained VT/VF during follow-up, with the input of significant variables from univariable logistic regression. (15,16) A multivariable logistic regression model was used as a benchmark for model comparison. Comparative analyses were conducted according to the area under the receiver operating characteristic curve (AUC, ROC), precision, recall, and F1 score. ROC curve, precision-recall curve, and lift curve were presented. Lift is a measure of the effectiveness of a predictive model calculated as the ratio between the results obtained with and without the predictive model. A lift curve is a way of visualizing the performance of a classification model. The greater the area between the lift curve and the baseline, the better the model. All analysis was performed using R Studio (Version: 1.3.1073).

## Results

### Baseline Characteristics and Predictors

The present cohort consists of 548 patients (7.3% females, age at diagnosis: 49.9±16.3 years old, follow-up duration: 84±55 months) (**Table 1**). In total, 66 patients experienced at least one episode of sustained VT/VF during follow-up. Only 9.7% of the cohort undergone genetic testing, and therefore these results were not included as predictors. Univariable Cox regression identified the following predictors of the primary outcome: 1) evolution of BrP (HR: 0.52, 95% CI: [0.29-0.94], p=0.030); 2) presentation of syncope (HR: 3.69, 95% CI: [2.08-6.58], p<0.0001); 3) other arrhythmia (HR: 2.89, 95% CI: [1.69-4.97], p<0.001); 4) early repolarization in peripheral leads (HR: 3.28, 95% CI: [1.62-6.64], p=0.001); 5) aVR sign (HR: 2.76, 95% CI: [1.43-5.34], p=0.003); 6) significant S wave in lead 1 (HR: 3.46, 95% CI: [1.96-6.11], p<0.0001); 7) QTc interval (HR: 1.01, 95% CI: [1.00-1.02], p=0.005); 8) initial VT/VF (HR: 9.67, 95% CI: [5.83-16.03], p<0.0001). Initial VT/VF was excluded as a multivariable predictor to avoid collinearity when accounting for disease manifestation. Syncope (HR: 3.03, 95% CI: [1.51, 6.07], p= 0.002), other arrhythmias (HR: 3.41, 95% CI: [1.76, 6.61], p<0.001) and significant S wave in lead 1 (HR: 2.15, 95% CI: [1.03, 4.50], p=0.042) remained significant on multivariable analysis (**Table 2**).

**Table 1.** Baseline and clinical characteristics of the study cohort of patients with Brugada syndrome.

| **Characteristics** | **Mean(SD);N or Count(%)** |
| --- | --- |
| **Clinical** |  |
| Female gender | 40(7.29%) |
| Age | 49.9(16.3) |
| Initial type 1 BrP | 340(62.04%) |
| Evolution of BrP | 187(34.12%) |
| Fever-induced BrP | 86(15.69%) |
| Drug challenge test | 234 (42.70%) |
| Positive drug challenge test | 204 (87.18%) |
| EPS | 112 (20.44%) |
| Positive EPS | 76 (67.86%) |
| Genetic test | 53 (9.67%) |
| Positive genetic test | 18 (33.96%) |
| Family history of BrS | 17(3.10%) |
| Family history of SCD | 45(8.21%) |
| Syncope | 236(43.06%) |
| VT/VF | 85(15.51%) |
| Initial VT/VF | 43(7.85%) |
| Other arrhythmias | 83(15.14%) |
| **Baseline electrocardiogram** |  |
| Early repolarization in peripheral leads | 39(7.11%) |
| aVR Sign | 55(10.03%) |
| Significant S wave in lead 1 | 101(18.43%) |
| Fragmented QRS | 62(11.31%) |
| Heart rate | 81.1(19.8);n=477 |
| PR interval | 169.0(28.7);n=463 |
| QRS interval | 105.8(22.2);n=475 |
| QTc Interval | 416.5(32.7);n=475 |
| QT interval | 368.2(41.7);n=475 |
| P wave axis | 61.7(22.6);n=463 |
| QRS axis | 59.4(40.1);n=472 |
| T wave axis | 54.7(26.2);n=473 |

BrP: Brugada electrocardiographic pattern; SCD: sudden cardiac death; EPS: electrophysiological study; VF: ventricular fibrillation; VT: ventricular tachycardia; BrS: Brugada Syndrome

**Table 2A.** Significant univariable and multivariable predictors of VT/VF during follow-up from Cox regression. * for *p* ≤ 0.05, ** for *p* ≤ 0.01, *** for *p* ≤ 0.001

| **Characteristics** | **Univariable**<br>**HR [95% CI];P value** | **Multivariable**<br>**HR [95% CI];P value** |
| --- | --- | --- |
| Male gender | 0.19[0.03-1.36];0.0985 | - |
| Female gender | 5.30[0.73-38.37];0.0985 | - |
| Age | 1.00[0.98-1.02];0.8602 | - |
| Initial type 1 BrP | 1.10[0.65-1.87];0.7301 | - |
| Evolution of BrP | 0.52[0.29-0.94];0.0303* | 0.55[0.27, 1.11];0.0948. |
| Fever-induced BrP | 0.48[0.17-1.34];0.1618 | - |
| Family history of BrS | 0.87[0.21-3.56];0.8409 | - |
| Family history of SCD | 1.10[0.44-2.75];0.8454 | - |
| Syncope | 3.69[2.08-6.58];<0.0001*** | 3.03[1.51, 6.07];0.0018** |
| Initial VT/VF | 9.67[5.83-16.03]; <0.0001*** | - |
| Other arrhythmia | 2.89[1.69-4.97];0.0001*** | 3.41[1.76, 6.61];0.0003*** |
| Early repolarization in peripheral leads | 3.28[1.62-6.64];0.0010** | 1.93[0.76, 4.90];0.1693 |
| aVR Sign | 2.76[1.43-5.34];0.0026** | 1.30[0.58, 2.90];0.5229 |
| Significant S wave in lead I | 3.46[1.96-6.11];<0.0001*** | 2.15[1.03, 4.50];0.0422* |
| Fragmented QRS | 1.75[0.86-3.58];0.1219 | - |
| Heart rate | 1.00[0.99-1.02];0.5128 | - |
| PR interval | 1.00[0.99-1.01];0.8199 | - |
| QRS interval | 1.00[0.99-1.01];0.8096 | - |
| QTc Interval | 1.01[1.00-1.02];0.0051** | 1.01[1.00, 1.02];0.1243 |
| QT interval | 1.01[1.00-1.01];0.0851 | - |
| P wave axis | 1.00[0.99-1.01];0.9914 | - |
| QRS axis | 1.00[0.99-1.01];0.9237 | - |
| T wave axis | 1.00[0.99-1.01];0.8246 | - |

BrP: Brugada electrocardiographic pattern; SCD: sudden cardiac death; VF: ventricular fibrillation; VT: ventricular tachycardia; BrS: Brugada Syndrome

**Table 2B.** Significant univariable and multivariable predictors of VT/VF during follow-up from logistic regression. * for *p* ≤ 0.05, ** for *p* ≤ 0.01, *** for *p* ≤ 0.001

| Characteristics | Odds Ratio [95%CI] | P-value |
| --- | --- | --- |
| Male gender | 0.37[0.09,1.55] | 0.1720 |
| Female gender | 2.74[0.65,11.63] | 0.1720 |
| Age | 1.0[0.98,1.01] | 0.6334 |
| <50 | 1.0[Reference] | - |
| 50-60 | 0.53[0.26,1.06] | 0.0743 |
| 60-70 | 0.93[0.47,1.86] | 0.8462 |
| 70-80 | 1.44[0.65,3.23] | 0.3696 |
| >=80 | 1.13[0.25,5.11] | 0.8764 |
| Initial type 1 BrP | 0.87[0.51,1.47] | 0.5983 |
| Evolution of BrP | 0.69[0.39,1.23] | 0.2125 |
| Fever-induced BrP | 0.41[0.16,1.04] | 0.0607 |
| Family history of BrS | 1.59[0.45,5.69] | 0.4746 |
| Family history of SCD | 0.91[0.34,2.38] | 0.8410 |
| Syncope | 3.53[2.03,6.16] | <0.0001 |
| VT/VF | - | - |
| Other arrhythmia | 2.89[1.61,5.21] | 0.0004*** |
| Early Repolarization in Peripheral Leads | 1.39[1.02,1.89] | 0.0379* |
| aVR Sign | 1.39[1.02,1.9] | 0.0376* |
| Significant S wave in Lead 1 | 1.73[1.27,2.35] | 0.0005*** |
| fragmented QRS | 1.32[0.96,1.8] | 0.0870 |
| Heart Rate | 1.0[0.99,1.01] | 0.6894 |
| PR interval | 1.0[0.99,1.01] | 0.8831 |
| QRS interval | 1.0[0.99,1.01] | 0.4507 |
| QTc Interval | 1.01[1.0,1.02] | 0.0123* |
| QT interval | 1.01[1.0,1.01] | 0.0852 |
| P wave axis | 0.99[0.98,1.0] | 0.1936 |
| QRS axis | 1.0[0.99,1.01] | 0.8595 |
| T wave axis | 0.98[0.97,0.99] | 0.0034** |

The importance ranking of the significant univariable predictors is displayed in **Supplementary Table 2**. Interestingly, the three significant multivariable predictors were not the three most important variables. Whilst significant S wave in lead 1 (minimal depth: 1.38) is ranked the most important, the occurrence of syncope (minimal depth: 2.25) and other arrhythmias (minimal depth: 2.23) were ranked lower on the list. The strength of the pairwise interactions amongst these variables is shown in **Figure 1**. QTc interval remains the most influential factor.

**Figure 1.**
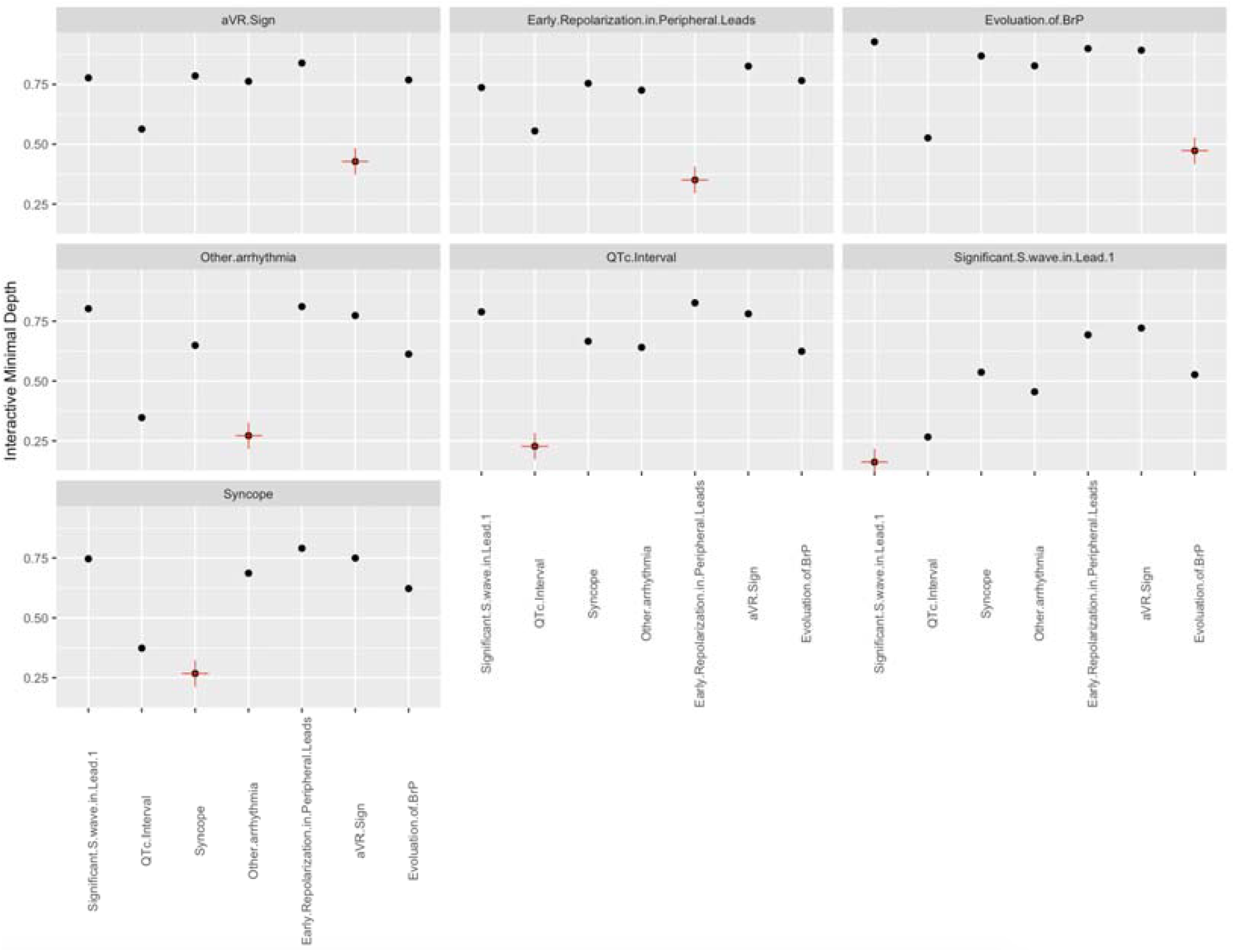
Pairwise interaction between significant univariable predictors.

### Performance of the existing scores and development of a new score

The score developed by Sieira *et al*. showed the best performance with an AUC of 0.805, followed by the Shanghai score (0.698), and the scores by Okamura *et al*. (0.667), Delise *et al*. (0.661), Letsas *et al*. (0.656) and Honarbakhsh *et al*. (0.592) (**Table 3**). A novel risk score was developed based on the following steps. Firstly, the best performing score with the highest AUC, was selected from the existing scores (the Sieira score). The original weighting of the Sieira score was used. Additional variables that were significant on univariable Cox regression were selected (arrhythmias other than ventricular tachyarrhythmias, ER pattern in the peripheral leads, aVR sign, S-wave in lead I, QTc ≥436 ms) (**Table 4**). This score has the highest AUC of 0.855 (95% CI: 0.808-0.901).

**Table 3.** Comparison of the performance of different scores.

| **Risk scores** | **AUC [95% CI] for whole cohort** | **AUC [95% CI] for intermediate risk group** |
| --- | --- | --- |
| New score | 0.855 [0.808-0.901] | 0.704 [0.580-0.828] |
| Sieira score | 0.805 [0.746-0.863] | 0.567 [0.412-0.721] |
| Shanghai score | 0.698 [0.623-0.772] | 0.490 [0.342-0.639] |
| Honarbakhsh score | 0.592 [0.513-0.671] | 0.452 [0.297-0.606] |
| Okamura score | 0.667 [0.600-0.733] | 0.543 [0.379-0.707] |
| Konstantinos score | 0.656 [0.591-0.721] | 0.483 [0.318-0.649] |
| Delise score | 0.661 [0.596-0.726] | 0.531 [0.382-0.682] |

**Table 4.** Novel risk score derived from the best performing score (Sieira score) and additional significant univariable predictors.

| **Characteristics** | **Score** |
| --- | --- |
| **Spontaneous type 1 Brugada pattern** | 1 |
| **Early familial SCD** | 1 |
| **Positive EPS** | 2 |
| **Syncope** | 2 |
| **Sinus node dysfunction** | 3 |
| **Prior aborted SCD** | 4 |
| **Other Arrhythmias (AT, AF, SVT)** | 1 |
| **ER pattern on peripheral leads** | 1 |
| **aVR sign** | 1 |
| **S-wave in lead I** | 1 |
| **QTc ≥ 436 ms** | 1 |

An intermediate risk subgroup was identified by ranking the patients based on our score into quartiles and including quartiles 2 and 3. All of the scores applied to this subgroup showed significantly lower AUCs. The newly developed score showed the best performance with an AUC of 0.704, followed by the scores by Sieira *et al*., Okamura *et al*., Delise *et al*., Shanghai score, Letsas *et al*. and Honarbakhsh *et al*. (**Table 3**).

### The Gaussian Naïve Bayes model and nearest neighbour model

Significant predictors identified on univariable logistic regression were used as input variables for the machine learning models, the nearest neighbor model and Gaussian Naïve Bayes model. Their ability to predict sustained VT/VF on follow-up was determined, with a five-fold cross validation approach. The multivariable logistic regression model was used as a benchmark for comparative analyses **(Figure 2)**. ROC curve, precision-recall curve, and lift curve are presented accordingly. Lift curve measures the effectiveness of a predictive model calculated as the ratio between the results obtained with and without the predictive model. A greater area between the lift curve of this model and that of the baseline model using multivariable logistic regression reflects a better model performance. The Gaussian Naïve Bayes model demonstrated the best performance with an AUC of 0.97, a F1 score of 0.87 and greatest area from the lift curve, compared to the logistic regression the nearest neighbor models (p for trends<0.001).

**Figure 2.**
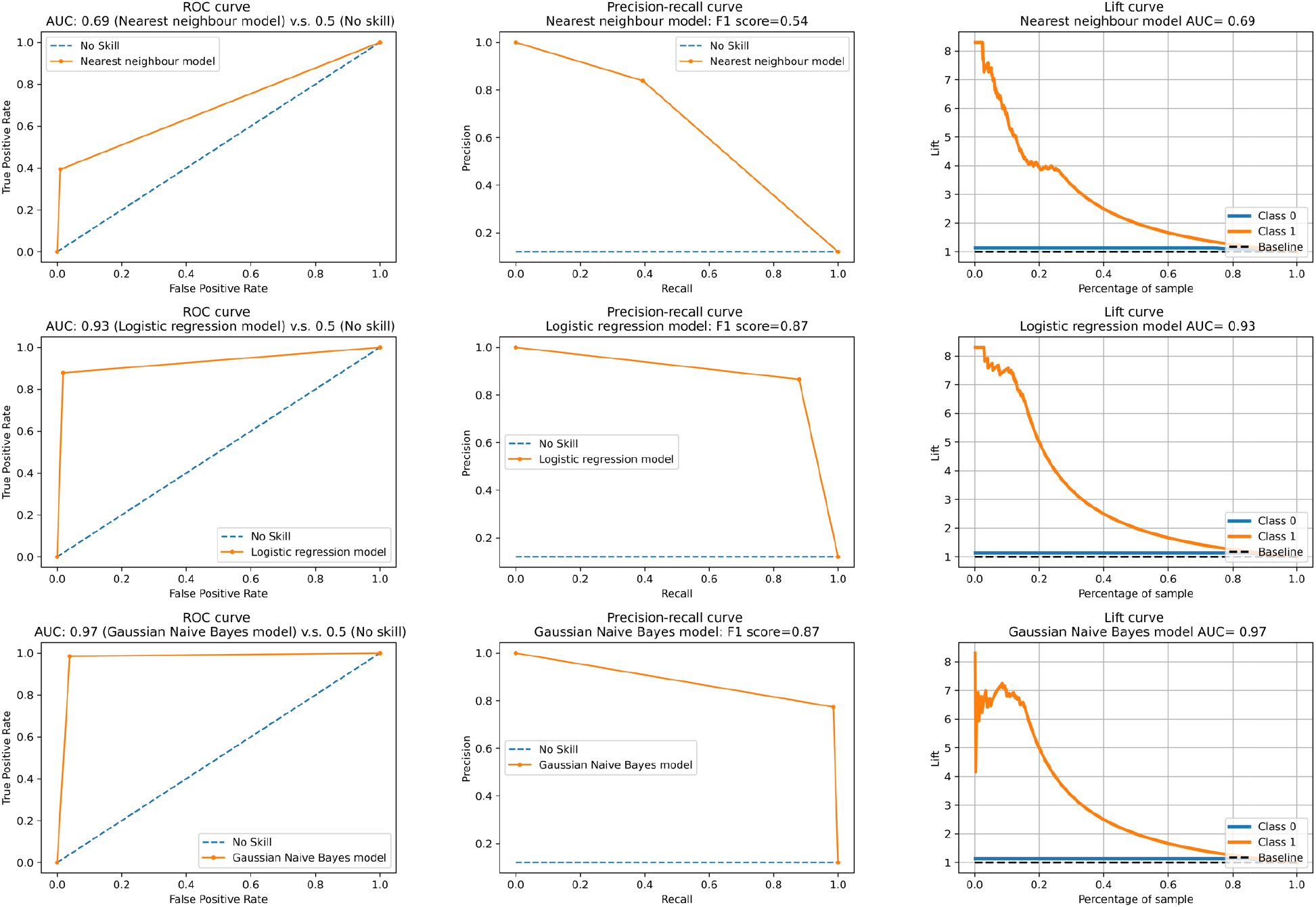
Performance comparisons of machine learning models to predict for sustained VT/VF during follow-up with significant univariable predictors as input variables.

## Discussion

To the best of our knowledge, this is the first Asian territory-wide BrS cohort study that directly compared all of the published risk scores. The major findings of the present study include: 1) simple multiparametric scores based on the combination of clinical and baseline electrocardiographic parameters can be used for risk stratification in BrS; 2) interactions between predictors can influence the predictive performance of the score; 3) spontaneous type 1 BrP, family history of SCD, syncope and inducible EPS can be useful for the risk stratification of intermediate risk patients.

Over the past decade, there has been increasing efforts in developing simple-to-use predictive scores for risk stratification in BrS. However, many either include findings from investigations that are only indicated for certain patient groups such as EPS, or include clinical or crude ECG parameters. (6,10-14) As a result, the scores are either difficult to be universally applied amongst all BrS patients, or have insufficient predictive power. Also, it should be noted that the Shanghai score was initially developed for a diagnostic, instead of a prognostic purpose. (12) The evidence supporting its use in risk stratification was based on demonstrations of differences in the arrhythmic events between patients with ≤3, 3.5, 4-5 and ≥5.5 points. (17) By contrast, Probst *et al*. found that whilst the Shanghai score had an AUC of 0.73 and was able to distinguish between extreme risk groups, it was unable to further stratify patients at intermediate risks (7).

The improved predictive performance of the novel risk score demonstrates that the inclusion of comprehensive clinical and baseline ECG indices is sufficient as an initial risk stratification tool that can be applied at the time of diagnosis.

Spontaneous type 1 BrP, family history of SCD, syncope and inducible arrhythmias detected during EPS are parameters common to the three predictive scores with the highest AUC in their prediction of intermediate risk patients. Since the manifestation of spontaneous type 1 BrP is a cornerstone of the diagnosis of BrS, the clinical presentation of the patient can range from asymptomatic to VT/VF. (18) Therefore, whilst patients who present with spontaneous type 1 BrP are considered to be at a higher risk, other clinical and electrocardiographic factors should be taken into account. (13,19,20) Similarly, the plethora of possible etiologies underlying syncope in BrS, ranging from benign causes to malignant arrhythmias, renders the need for multiparametric assessment in patients presenting with syncope. (21) By contrast, a family history of SCD is a marker for intermediate risk is likely due to the polygenic inheritance, variable expression and incomplete penetrance in BrS. (22) Whilst the presence of pathogenic SCN5A mutation increases the risk of BrS manifestation, clinical and environmental factors are required to drive the degree of electrophysiological dysfunction across the disease threshold. (23,24) Although genetic data were not examined in the present study, the predictive value of genetic and genomic findings should be explored in the future (25).

The prognostic value of EPS remains controversial. The current evidence on the predictive power is mixed, varying between different patient subgroups. (10,26,27) A recent meta-analysis suggests that its risk stratification value is operator- and protocol-dependent. (28) Therefore, EPS should be applied on an individual basis with particular considerations towards patient factors, using standardized protocols with predefined locations for the placement of stimulation electrodes and the pacing protocols. It may be useful for particular subgroups of patients, for example, prior studies have reported that in the case of syncope of unknown etiology, the presence of inducible EPS may reflect a higher SCD risk. (29,30)

### Limitations

Several limitations for the present study should be noted. Firstly, due to the limitations in the availability of certain variables needed for particular risk scores, these scores could not be fully applied to the present cohort. For example, nocturnal agonal respiration and family history to second-degree relatives were not recorded in case notes, and thus only a limited version of the Shanghai score was calculated. Secondly, the etiology of syncope was not documented, thus syncope of non-arrhythmic origin may be included. Thirdly, given the low rates of EPS and genetic test performance, the predictive value of findings from these two tests was not assessed. Finally, our score does not incorporate latent interactions between the risk variables, which have previously been shown to be important for risk stratification (31,32). Future studies with the integration of machine learning techniques into the predictive scores may improve the accuracy of risk stratification through the recognition of latent interactions between predictors.

## Conclusion

In conclusion, simple risk scores consisting of clinical and baseline electrocardiographic indices are useful in the risk stratification of the overall BrS population. However, the inclusion of investigation results and more complex models are needed to improve the predictive performance of risk scores against the intermediate risk BrS population. The incorporation of machine learning and genomics may be a direction for future research to improve the stratification of SCD risk amongst BrS patients.

## Data Availability

All data produced in the present study are available upon reasonable request to the authors. A deidentified dataset has been made available at https://zenodo.org/record/4383364

## References

1. Probst V, Veltmann C, Eckardt L et al. Long-term prognosis of patients diagnosed with Brugada syndrome: Results from the FINGER Brugada Syndrome Registry. Circulation 2010;121:635–43.

2. Michowitz Y, Milman A, Andorin A et al. Characterization and Management of Arrhythmic Events in Young Patients With Brugada Syndrome. J Am Coll Cardiol 2019;73:1756–1765.

3. Vutthikraivit W, Rattanawong P, Putthapiban P et al. Worldwide Prevalence of Brugada Syndrome: A Systematic Review and Meta-Analysis. Acta Cardiol Sin 2018;34:267–277.

4. Milman A, Hochstadt A, Andorin A et al. Time-to-first appropriate shock in patients implanted prophylactically with an implantable cardioverter-defibrillator: data from the Survey on Arrhythmic Events in BRUgada Syndrome (SABRUS). Europace 2019;21:796–802.

5. Priori SG, Gasparini M, Napolitano C et al. Risk stratification in Brugada syndrome: results of the PRELUDE (PRogrammed ELectrical stimUlation preDictive valuE) registry. J Am Coll Cardiol 2012;59:37–45.

6. Sieira J, Conte G, Ciconte G et al. A score model to predict risk of events in patients with Brugada Syndrome. Eur Heart J 2017;38:1756–1763.

7. Probst V, Goronflot T, Anys S et al. Robustness and relevance of predictive score in sudden cardiac death for patients with Brugada syndrome. Eur Heart J 2020.

8. Letsas KP, Asvestas D, Baranchuk A et al. Prognosis, risk stratification, and management of asymptomatic individuals with Brugada syndrome: A systematic review. Pacing Clin Electrophysiol 2017;40:1332–1345.

9. Antzelevitch C, Yan GX, Ackerman MJ et al. J-Wave syndromes expert consensus conference report: Emerging concepts and gaps in knowledge. Europace 2017;19:665–694.

10. Letsas KP, Bazoukis G, Efremidis M et al. Clinical characteristics and long-term clinical course of patients with Brugada syndrome without previous cardiac arrest: a multiparametric risk stratification approach. Europace 2019;21:1911–1918.

11. Honarbakhsh S, Providencia R, Garcia-Hernandez J et al. A Primary Prevention Clinical Risk Score Model for Patients With Brugada Syndrome (BRUGADA-RISK). JACC Clin Electrophysiol 2021;7:210–222.

12. Antzelevitch C, Yan GX, Ackerman MJ et al. J-Wave syndromes expert consensus conference report: Emerging concepts and gaps in knowledge. Heart Rhythm 2016;13:e295–324.

13. Delise P, Allocca G, Marras E et al. Risk stratification in individuals with the Brugada type 1 ECG pattern without previous cardiac arrest: usefulness of a combined clinical and electrophysiologic approach. Eur Heart J 2011;32:169–76.

14. Okamura H, Kamakura T, Morita H et al. Risk stratification in patients with Brugada syndrome without previous cardiac arrest - prognostic value of combined risk factors. Circ J 2015;79:310–7.

15. Peter Hall BUP, Richard J. Samworth. Choice of neighbor order in nearest-neighbor classification. Annals of Statistics 2009.

16. Daniele Soria JMG, Federico Ambrogi, Elia M. Biganzoli, Ian O. Ellis. A ‘non-parametric’ version of the naive Bayes classifier. Knowledge-Based Systems 2011.

17. Kawada S, Morita H, Antzelevitch C et al. Shanghai Score System for Diagnosis of Brugada Syndrome: Validation of the Score System and System and Reclassification of the Patients. JACC Clin Electrophysiol 2018;4:724–730.

18. Brugada J, Campuzano O, Arbelo E, Sarquella-Brugada G, Brugada R. Present Status of Brugada Syndrome: JACC State-of-the-Art Review. J Am Coll Cardiol 2018;72:1046–1059.

19. Miyamoto A, Hayashi H, Makiyama T et al. Risk determinants in individuals with a spontaneous type 1 Brugada ECG. Circ J 2011;75:844–51.

20. Asvestas D, Tse G, Baranchuk A et al. High risk electrocardiographic markers in Brugada syndrome. Int J Cardiol Heart Vasc 2018;18:58–64.

21. Mascia G, Bona RD, Ameri P et al. Brugada syndrome and syncope: a practical approach for diagnosis and treatment. Europace 2020.

22. Rowe MK, Roberts JD. The evolution of gene-guided management of inherited arrhythmia syndromes: Peering beyond monogenic paradigms towards comprehensive genomic risk scores. J Cardiovasc Electrophysiol 2020;31:2998–3008.

23. Bezzina CR, Barc J, Mizusawa Y et al. Common variants at SCN5A-SCN10A and HEY2 are associated with Brugada syndrome, a rare disease with high risk of sudden cardiac death. Nat Genet 2013;45:1044–9.

24. Lek M, Karczewski KJ, Minikel EV et al. Analysis of protein-coding genetic variation in 60,706 humans. Nature 2016;536:285–91.

25. Zhang Z-H, Barajas-Martínez H, Xia H et al. Distinct Features of Probands With Early Repolarization and Brugada Syndromes Carrying SCN5A Pathogenic Variants. Journal of the American College of Cardiology 2021;78:1603–1617.

26. Li X, Sacher F, Kusano KF et al. Pooled Analysis of Risk Stratification of Spontaneous Type 1 Brugada ECG: Focus on the Influence of Gender and EPS. Front Physiol 2018;9:1951.

27. Rodriguez-Manero M, Jorda P, Hernandez J et al. Long-term prognosis of women with Brugada syndrome and electrophysiological study. Heart Rhythm 2020.

28. Sroubek J, Probst V, Mazzanti A et al. Programmed Ventricular Stimulation for Risk Stratification in the Brugada Syndrome: A Pooled Analysis. Circulation 2016;133:622–30.

29. Hernandez-Ojeda J, Arbelo E, Jorda P et al. The role of clinical assessment and electrophysiology study in Brugada syndrome patients with syncope. Am Heart J 2020;220:213–223.

30. Giustetto C, Cerrato N, Ruffino E et al. Etiological diagnosis, prognostic significance and role of electrophysiological study in patients with Brugada ECG and syncope. Int J Cardiol 2017;241:188–193.

31. Lee S, Zhou J, Li KHC et al. Territory-wide cohort study of Brugada syndrome in Hong Kong: predictors of long-term outcomes using random survival forests and non-negative matrix factorisation. Open Heart 2021;8.

32. Tse G, Zhou J, Lee S et al. Incorporating Latent Variables Using Nonnegative Matrix Factorization Improves Risk Stratification in Brugada Syndrome. J Am Heart Assoc 2020:e012714.

