## Supplementary Appendix for "Comparing the performance of risk stratification scores in Brugada syndrome: a multi-centre study"

**Supplementary Table 1. Existing predictive scores for risk stratification in Brugada syndrome.**

| Score | Variable | Weight |
|---|---|---|
| Sieira score | Spontaneous type 1 BrP | 1 |
| | Early familial SCD | 1 |
| | Inducible EPS | 2 |
| | Syncope | 2 |
| | Sinus node dysfunction | 3 |
| | SCD | 4 |
| Shanghai score | ECG: | |
| | • Spontaneous type 1 BrP at high* or nominal leads | 3.5 |
| | • Fever-induced type 1 BrP at high* or nominal leads | 3 |
| | • Type 2 or 3 BrP converting with drug challenge | 2 |
| | Clinical history: | |
| | • Unexplained cardiac arrest or documented VF/ polymorphic VT | 3 |
| | • Nocturnal agonal respiration* | 2 |
| | • Suspected arrhythmic syncope* | 2 |
| | • Syncope of unclear etiology | 1 |
| | • Atrial flutter/ fibrillation in patients < 30 years old without alternative etiology | 0.5 |
| | Family history: | |
| | • First/ second degree* relative with definite BrS | 2 |
| | • Suspected SCD in first/ second degree* relative | 1 |
| | • Unexplained SCD < 45 years old in first/ second degree * relative with negative biopsy | 0.5 |
| | Genetic test: | |
| | • Probable pathogenic mutation in BrS susceptibility gene | 0.5 |
| Honarbakhsh score | Probable arrhythmia-related syncope* | 12 |
| | Spontaneous type 1 BrP | 14 |
| | Early repolarization in peripheral leads | 9 |
| | Type 1 BrP in peripheral leads | 12 |

| | | |
|---|---|---|
| Okamura score | Spontaneous type 1 BrP | 1 |
| | Syncope | 1 |
| | Inducible EPS | 1 |
| Konstantinos score | Spontaneous type 1 BrP | 1 |
| | Syncope | 1 |
| | Family history of SCD | 1 |
| | Fragmented-QRS | 1 |
| | QRS duration in lead V2* | 1 |
| Delise score | Spontaneous type 1 BrP | 1 |
| | Syncope | 1 |
| | Family history of SCD | 1 |

BrP: Brugada electrocardiographic pattern; SCD: sudden cardiac death; EPS: electrophysiological study; VF: ventricular fibrillation; VT: ventricular tachycardia; BrS: Brugada Syndrome

**Supplementary Table 2. Importance ranking of variables using random survival forest model**

| Characteristics | Minimal Depth | Variable Importance |
|---|---|---|
| Significant S wave in Lead 1 | 1.38 | 0.09529 |
| QTc Interval | 1.85 | 0.00915 |
| Other arrhythmias | 2.23 | 0.03002 |
| Syncope | 2.25 | 0.01165 |
| Early Repolarization in Peripheral Leads | 2.80 | 0.01121 |
| aVR Sign | 3.43 | 0.00430 |

Other arrhythmias are defined as sick sinus syndrome, bradycardia, atrioventricular block, supraventricular tachycardia (SVT), supraventricular ectopic beats (SVE), atrial fibrillation (AF), atrial tachycardia (AT), or atrial flutter.
